# Rates of COVID-19-related Outcomes in Cancer compared to non-Cancer Patients

**DOI:** 10.1101/2020.08.14.20174961

**Authors:** Lova Sun, Sanjna Surya, Anh N. Le, Heena Desai, Abigail Doucette, Peter Gabriel, Marylyn Ritchie, Daniel Rader, Ivan Maillard, Erin Bange, Alexander Huang, Robert H. Vonderheide, Angela DeMichele, Anurag Verma, Ronac Mamtani, Kara N. Maxwell

## Abstract

Cancer patients are a vulnerable population postulated to be at higher risk for severe COVID-19 infection. Increased COVID-19 morbidity and mortality in cancer patients may be attributable to age, comorbidities, smoking, healthcare exposure, and cancer treatments, and partially to the cancer itself. Most studies to date have focused on hospitalized patients with severe COVID-19, thereby limiting the generalizability and interpretability of the association between cancer and COVID-19 severity. We compared outcomes of SARS-CoV-2 infection in 323 patients enrolled prior to the pandemic in a large academic biobank (n=67 cancer patients and n=256 non-cancer patients). After adjusting for demographics, smoking status, and comorbidities, a diagnosis of cancer was independently associated with higher odds of hospitalization (OR 2.16, 95% CI 1.12-4.18) and 30-day mortality (OR 5.67, CI 1.49-21.59). These associations were primarily driven by patients with active cancer. These results emphasize the critical importance of preventing SARS-CoV-2 exposure and mitigating infection in cancer patients.

---

Cancer patients appear to have a higher risk of COVID-19-related complications and mortality compared to the general population [1-10], due in part to factors such as older age, higher smoking rates, more comorbidities, frequent exposure to health care settings, and immunomodulatory effects of cancer therapies. In a multicenter study from Wuhan, China, patients with cancer hospitalized with COVID-19 infection were found to have higher rates of ICU admission, invasive ventilation, and severe symptoms when compared with age-matched non-cancer controls [9]. Similarly, in a New York City hospital system, admitted cancer patients were found to have higher risk of severe COVID-19 compared to non-cancer patients matched for age, sex, and comorbidities [11]. Additionally, recent administration of anti-cancer therapies has been associated with higher risk of mortality or complications from SARS-CoV-2 [7-12]. Because most studies have focused on cancer patients hospitalized with severe COVID-19, it is unclear whether cancer status has an independent adverse impact on clinical outcomes in a health system population-based group of patients diagnosed with SARS-CoV-2 infection. We leveraged the Penn Medicine Biobank (PMBB) at the University of Pennsylvania, an academic biobank allowing access to electronic health record (EHR) data [13], to investigate whether patients with cancer had worse COVID-19 outcomes than non-cancer patients.

Patients had previously consented to enrollment in PMBB prior to the onset of the COVID-19 pandemic, and were subsequently found to have SARS-CoV-2 infection by reverse transcriptase polymerase chain reaction (RT-PCR). Patients were defined as having a cancer diagnosis if they met at least one of three criteria: 1) three ICD-10 billing codes for an invasive (non-secondary) cancer, 2) inclusion in the Penn Medicine Cancer Registry, 3) one visit within a cancer service line clinic. All cancer diagnoses were confirmed by manual chart review. Patient characteristics and clinical outcomes (hospitalization, ICU admission, and 30-day mortality) were extracted from the EHR and compared in patients with and without cancer. Separate multivariable logistic regressions were performed to estimate odds ratios (OR) and 95% confidence intervals (CI) for the association between cancer diagnosis and COVID-19 outcomes (hospitalization, ICU admission, and mortality in the 30 days following COVID-19 diagnosis), adjusted for potential confounders including demographic factors, smoking status, comorbidities, and socioeconomic status estimated by the national poverty index based on neighborhood mapping [14, 15]. Exploratory subgroup analyses were performed to investigate these associations among patients with active cancer (defined as either having metastatic disease and/or receiving cancer-directed systemic therapy, radiation therapy, or surgical resection in the two months prior to COVID-19 diagnosis) compared to non-cancer patients, as well as those with cancer in remission compared to non-cancer patients.

As of June 2020, of 4,816 patients previously enrolled in PMBB who had been tested for COVID-19, 323 (7.3%) had laboratory-confirmed SARS-CoV-2 infection. Of COVID-19-positive patients, 67 (20.7%) had a cancer diagnosis (80.6% with solid tumor malignancy; and 26.9% with active cancer). Compared to non-cancer patients, COVID-19-positive cancer patients were more likely to be older (62 vs 50 years, p<0.001), male (53.7% vs 39.5%, p=0.035), and have a history of smoking (55.2% vs 35.%, p=0.003, **Table 1**). Notably, the proportion of Black patients was significantly higher in both cancer and non-cancer COVID-19-positive patients (65.7% and 64.1%, respectively), compared to all PMBB patients tested for SARS-CoV-2 (32.0%, p<0.001).

**Table 1.** Baseline Characteristics. Cancer vs Non-Cancer COVID-19-Positive Patients.

|  | Cancer (N=67) | No cancer (N=256) | p-value* |
| --- | --- | --- | --- |
| Age | 62 (53-71) | 50 (37-60) | <0.001 |
| Race |  |  |  |
| White | 18 (26.9%) | 74 (28.9%) | 0.980 |
| Black | 44 (65.7%) | 164 (64.1%) |  |
| Asian | 2 ( 3.0%) | 5 ( 2.0%) |  |
| Other | 2 ( 3.0%) | 8 ( 3.1%) |  |
| Unknown | 1 ( 1.5%) | 5 ( 2.0%) |  |
| Gender |  |  |  |
| Female | 31 (46.3%) | 155 (60.5%) | 0.035 |
| Male | 36 (53.7%) | 101 (39.5%) |  |
| Ever Smoker | 37 (55.2%) | 90 (35.2%) | 0.003 |
| National Poverty Percentile | 91 (63-93) | 91 (76.5-93) | 0.290 |
| Comorbidities |  |  |  |
| Hypertension | 38 (56.7%) | 117 (45.7%) | 0.110 |
| Diabetes | 22 (32.8%) | 72 (28.1%) | 0.450 |
| Obesity | 12 (17.9%) | 76 (29.7%) | 0.054 |
| Pulmonary disease | 14 (20.9%) | 36 (14.1%) | 0.170 |
| Mood disorders | 8 (11.9%) | 37 (14.5%) | 0.600 |
| Ischemic cardiovascular disease | 11 (16.4%) | 29 (11.3%) | 0.260 |
| Immunodeficiency | 5 ( 7.5%) | 11 ( 4.3%) | 0.290 |
| Cerebrovascular disease | 3 ( 4.5%) | 8 ( 3.1%) | 0.590 |

\*Wilcoxon rank-sum test for continuous outcomes; Pearson's chi-squared test for categorical outcomes

Rates of hospitalization, ICU admission, and 30-day mortality were higher in patients with cancer compared to those without cancer (55.2% vs 28.9%, 25.4% vs 11.7%, 13.4% vs 1.6%, respectively) (**Table 2**). Older age, Black race, and number of comorbidities were significantly associated with increased odds of hospitalization and ICU admission (all p<0.05). In fully adjusted models, cancer diagnosis was associated with statistically significantly increased odds of hospitalization (OR=2.16, 95% CI 1.12-4.18) and 30-day mortality (OR=5.67, 95% CI 1.49-21.59), and trend toward increased odds of ICU admission (OR=1.91, 95% CI 0.90-4.06). In exploratory subgroup analyses by cancer status (i.e. active vs. remission), adjusted associations with hospitalization, ICU admission, and 30-day mortality were strongest in the subgroup of patients with active cancer compared to non-cancer patients (adjusted OR 4.25, 3.28, and 28.8 respectively, all p<0.01), suggesting that the association between cancer diagnosis and poor COVID-19 outcomes was driven primarily by patients with active cancer.

**Table 2.**
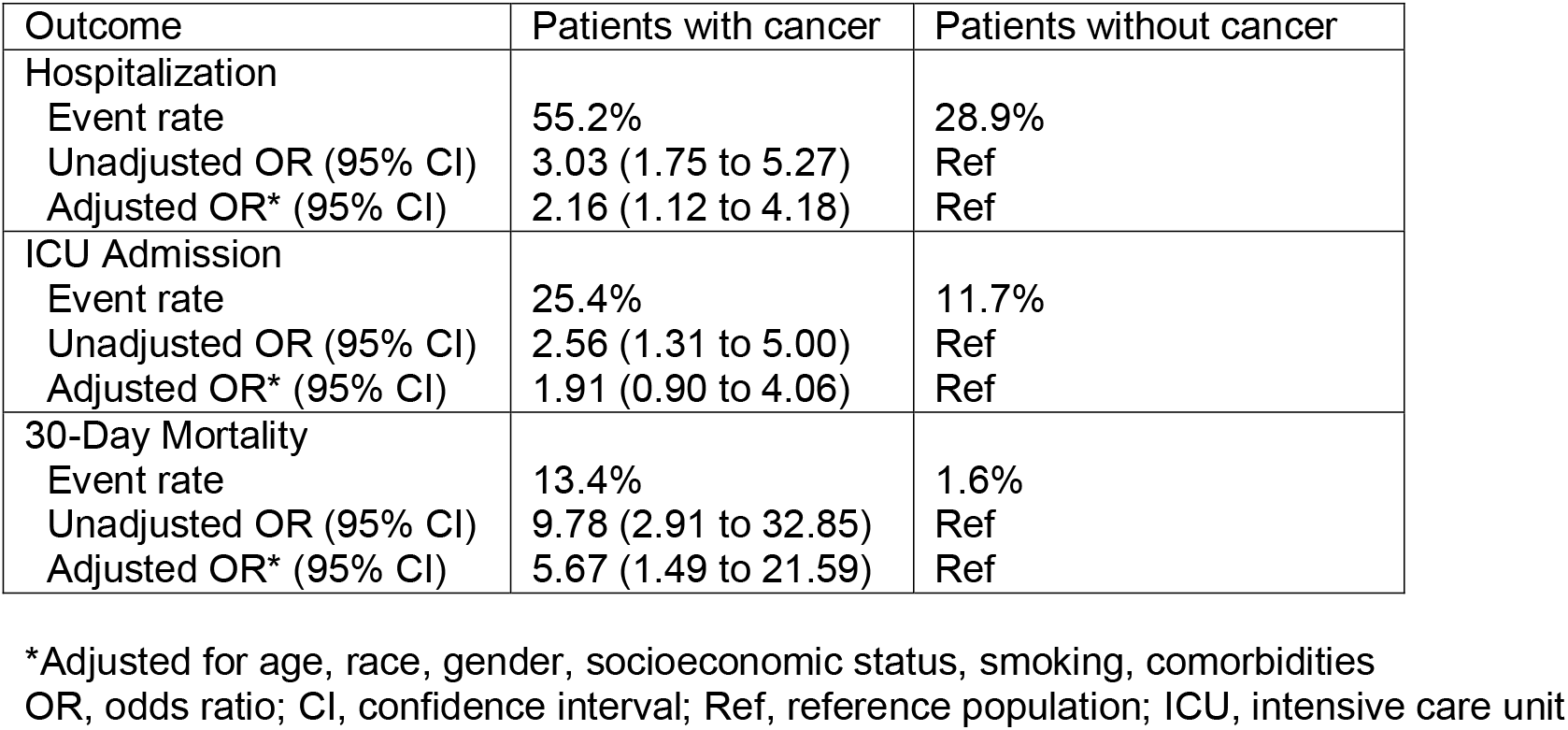
Event Rates and Odds Ratios of Clinical Outcome in Cancer vs Non-Cancer COVID-19-Positive Patients.

Concerns about risk of COVID-19 in cancer patients have led to alterations in cancer care such as treatment regimen modifications, delayed screening, decreased clinical trial enrollment, and increased telemedicine use [16] - the potential effects of which on cancer outcomes are as of yet unknown, although models predict a negative impact on survival [17]. Prior studies investigating the association between cancer and COVID-19 outcomes have largely focused on hospitalized patients with severe COVID-19 disease, who may not be representative of the general population. In particular, studies utilizing non-random samples, of which hospitalized patients are a classic example, are prone to collider bias, which can induce spurious or distorted associations [18]. We studied COVID-19-positive patients identified from a large pre-existing biobank, which allowed more unbiased population sampling of cancer and non-cancer patients. Our finding that cancer patients with COVID-19 were more likely than non-cancer patients to experience hospitalization and death even after adjusting for patient-level factors supports the hypothesis that cancer is an independent risk factor for poor COVID-19 outcomes.

Our study had several limitations. Our sample size did not allow detailed subgroup analysis of different cancer types, which may confer non-uniform risk of severe COVID-19 [1, 5, 8]. Similarly, our cohort contained a relatively small number of patients with active cancer, which precluded analysis of the impact of different cancer therapies on COVID-19 outcomes. A prospective observational study of patients in a United Kingdom cancer center network did not find an association between cancer therapy type in the past four weeks and mortality [19]. Given the conflicting evidence on the impacts of active vs. non-active cancer status, cancer stage, and cancer therapies on COVID-19 prognosis, deeper investigation of these variables is needed [2, 5, 9, 10, 12, 19].

In conclusion, in our cohort of COVID-19-positive patients identified from a health system population-based academic biobank, a diagnosis of cancer was strongly and independently associated with poor COVID-19 clinical outcomes including hospitalization and 30-day mortality. Given that it is critically important that these patients engage with the healthcare system for optimal cancer-directed management, our results suggest that patients with cancer, particularly those receiving active treatment, should be among groups specifically targeted for COVID-19 mitigation and prevention strategies such as vaccination.

## Data Availability

The data that support the findings of this study are available on request and subject to data use agreement. The data are not publicly available due to information that could compromise the privacy of research participants.

**Abbreviations**
COVID-19: Coronavirus disease 2019
SARS-CoV-2: severe acute respiratory syndrome coronavirus 2
EHR: electronic health records
ICU: intensive care unit
OR: odds ratio
CI: confidence interval
PMBB: Penn Medicine Biobank
Ref: reference population

## Funding

This work was supported by the National Institutes of Health (P30-CA016520 to Abramson Cancer Center)

Notes

*Role of Funder:* Salary support

*Author Disclosures:* Dr. Vonderheide reports having received consulting fees or honoraria from Celldex, Lilly, Medimmune, and Verastem; and research funding from Apexigen, Fibrogen, Inovio, Janssen, and Lilly. He is an inventor on a licensed patent relating to cancer cellular immunotherapy and receives royalties from Children’s Hospital Boston for a licensed research-only monoclonal antibody. Other authors declare that they have no competing interests.

## Acknowledgements

We acknowledge the Penn Medicine BioBank (PMBB) for providing data and thank the patient-participants of Penn Medicine who consented to participate in this research program. The PMBB is approved under IRB protocol# 813913 and supported by Perelman School of Medicine at University of Pennsylvania.

